# Nationwide Effectiveness of First and Second SARS-CoV2 Booster Vaccines during the Delta and Omicron Pandemic Waves in Hungary (HUN-VE 2 Study)

**DOI:** 10.1101/2022.03.27.22273000

**Authors:** Zoltán Kiss, István Wittmann, Lőrinc Polivka, György Surján, Orsolya Surján, Zsófia Barcza, Gergő Attila Molnár, Dávid Nagy, Veronika Müller, Krisztina Bogos, Péter Nagy, István Kenessey, András Wéber, Mihály Pálosi, János Szlávik, Zsuzsa Schaff, Zoltán Szekanecz, Cecília Müller, Miklós Kásler, Zoltán Vokó

## Abstract

**Background:** In Hungary, the pandemic waves in late 2021 and early 2022 were dominated by the Delta and Omicron SARS-CoV-2 variants, respectively. Booster vaccines were offered with one or two doses for the vulnerable population during these periods.

**Methods and Findings:** The nationwide HUN-VE 2 study examined the effectiveness of primary immunization, single booster, and double booster vaccination in the prevention of Covid-19 related mortality during the Delta and Omicron waves, compared to an unvaccinated control population without prior SARS-CoV-2 infection during the same study periods.

The risk of Covid-19 related death was 55% lower during the Omicron vs. Delta wave in the whole study population (n=9,569,648 and n=9,581,927, respectively; rate ratio [RR]: 0.45, 95% confidence interval [CI]: 0.44–0.48). During the Delta wave, the risk of Covid-19 related death was 74% lower in the primary immunized population (RR: 0.26; 95% CI: 0.25–0.28) and 96% lower in the booster immunized population (RR: 0.04; 95% CI: 0.04–0.05), vs. the unvaccinated control group. During the Omicron wave, the risk of Covid-19 related death was 40% lower in the primary immunized population (RR: 0.60; 95% CI: 0.55–0.65) and 82% lower in the booster immunized population (RR: 0.18; 95% CI: 0.16–0.2) vs. the unvaccinated control group. The double booster immunized population had a 93% lower risk of Covid-19 related death compared to those with only one booster dose (RR: 0.07; 95% CI. 0.01–0.46). The benefit of the second booster was slightly more pronounced in older age groups.

**Conclusions:** The HUN-VE 2 study demonstrated the significantly lower risk of Covid-19 related mortality associated with the Omicron vs. Delta variant and confirmed the benefit of single and double booster vaccination against Covid-19 related death. Furthermore, the results showed the additional benefit of a second booster dose in terms of SARS-CoV-2 infection and Covid-19 related mortality.

## 1 Introduction

The first coronavirus disease 2019 (Covid-19) cases were confirmed more than 2 years ago, and several new variants of the SARS-CoV-19 virus have been identified since then (1). Vaccines against SARS-CoV-19 infection with various mechanisms of action became available at the end of 2020 (2–4). In Hungary, 6 different vaccine types were approved by local regulatory authorities during the first half of 2021, when the B.1.1.7 (Alpha) variant dominated the pandemic wave. The HUN-VE study was published in the second half of 2021 and reported high or very high effectiveness for 5 different vaccine types against SARS-CoV-19 infection and Covid-19 related mortality caused by the alpha variant (3). In the meantime, several studies reported declining vaccine effectiveness 6–9 months after the first two doses (5–8). Moreover, the emergence of the new, more contagious Delta variant (B.1.617.2) led to new pandemic waves during the second half of 2021. Therefore, according to the recommendations by Israeli scientists, booster doses of different Covid-19 vaccines became available in several countries (9,10). Since August 1, 2021, the Hungarian government recommends heterologous or homologous booster vaccination at least 4 months after the primary vaccination course, particularly for vulnerable patient populations including those aged 60 or older and people with chronic diseases (11). The effectiveness of booster vaccines against SARS-CoV-19 infection after waning vaccine immunity was rapidly demonstrated by a number of studies, however, few analyses reported benefits regarding severe outcomes and death (9,10). By the end of 2021, the new Omicron variant reached Europe, which led to the highest-ever daily infection rates in early 2022 with lower mortality rates (12,13). In the meantime, certain countries such as Hungary started recommending a second booster shot for the vulnerable population including people aged 60 years or older and those having chronic diseases, however, the fourth dose was available for the whole population (14–16).

The aim of the HUN-VE 2 study was to examine the effectiveness of primary immunization as well as first and second booster doses during the Delta and Omicron waves of the SARS-CoV-2 pandemic in Hungary.

## 2 Methods

This nationwide, retrospective, observational study examined the overall effectiveness of primary immunization (first two vaccine doses, or one dose of the Janssen vaccine), first, and second booster vaccines against Covid-19 related death during ascending phases of the Delta and Omicron waves using data from the Hungarian Coronavirus Registry owned by the National Public Health Center (NPHC). We examined two 54-day long observation periods for the outcome of Covid-19 related death: 8 November 2021 to 31 December 2021 (dominant variant: Delta); 1 January 2022 to 23 February 2022 (dominant variant: Omicron) (Figure 1) (14).

**Figure 1.**
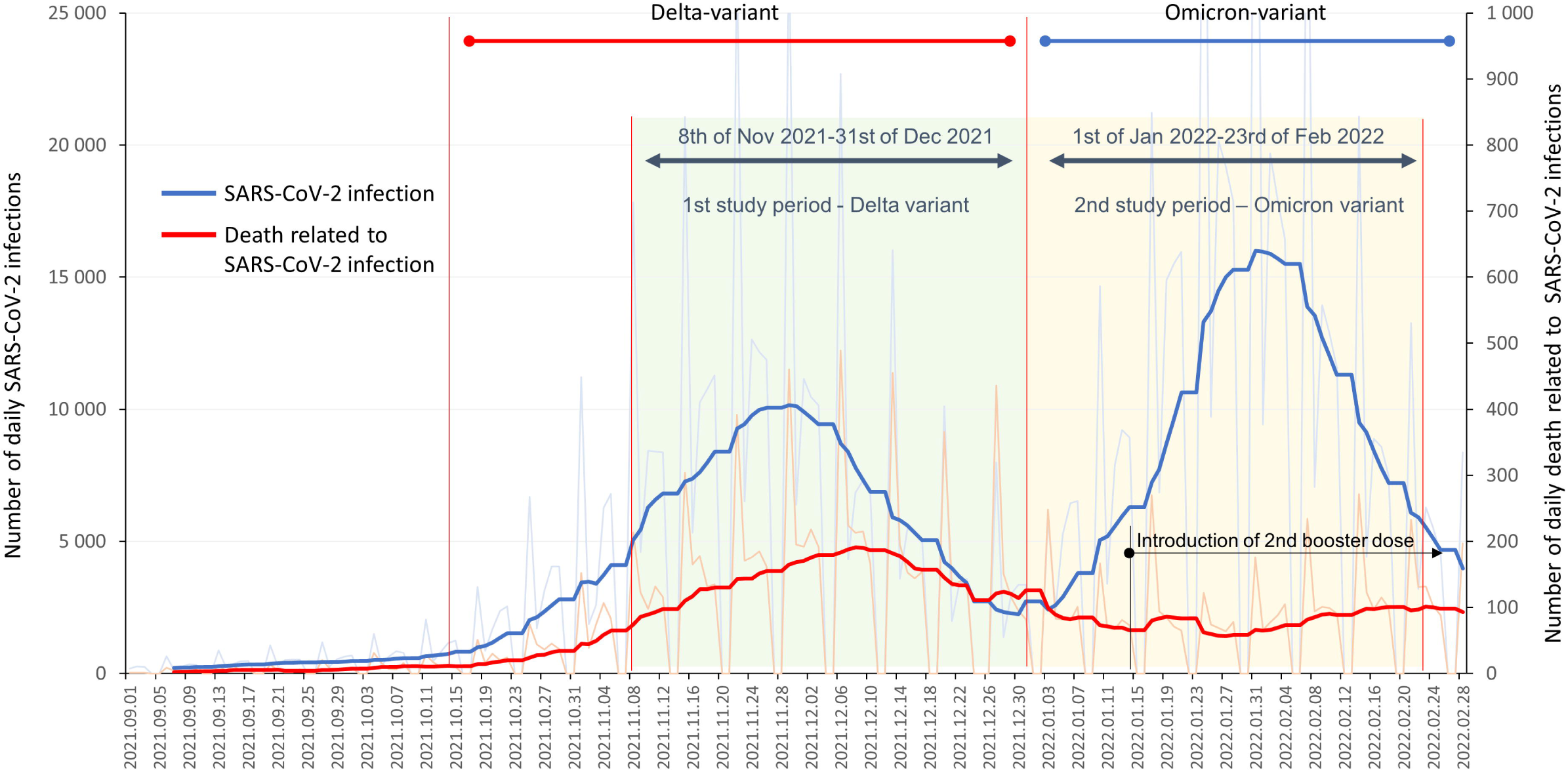
Study periods and timeline of the HUN-VE 2 study together with the reported daily SARS-CoV-2 infections and Covid-19 related mortality in Hungary between 1 September 2021, and 28 February 2022 (source: koronavirus.gov.hu).

Our study covers almost 100% of the Hungarian population, aged 16 years or older. The study included Hungarian residents who had active health insurance on 3 March 2021 and were alive on the first day of the respective study period. Cases of SARS-CoV-2 infection were reported by hospital physicians and general practitioners, collected by the NPHC on a daily basis using the centralized system of the National Social Information System (NSIS), and recorded in the Hungarian Coronavirus Register. Covid-19 related mortality was defined as death during SARS-CoV-2 positivity, regardless of whether death was the direct consequence of SARS-CoV-2 infection or other underlying causes (except for clear alternative causes such as trauma). The definition was the same used in the HUN-VE study (3).

Six different vaccine types and their combinations were available in Hungary prior to and during the study period: Pfizer-BioNTech, Moderna, AstraZeneca, Sputnik-V, Sinopharm, Janssen. The separate evaluation of vaccine effectiveness and different vaccine combinations will be reported by a subsequent HUN-VE study.

On any day, individuals without any record of SARS-Cov-2 infection at the beginning of each study period were classified as having completed primary immunization if at least 14 days had passed since the administration of the second dose of any vaccine type (or one dose of the Janssen vaccine), booster immunized starting from 14 days after the third dose (or second after the Janssen vaccine), and double booster immunized if at least 14 days had passed since the second booster dose (four times vaccinated or triple vaccinated in the case of the Janssen vaccine). The unvaccinated, control population included individuals who had not received any dose of any Covid-19 vaccine type and had no record of SARS-CoV-2 infection in the Hungarian Coronavirus Register, had active health insurance on 3 March 2021, and were alive on the first day of the respective study period.

The distribution of different types of vaccines as primary immunization, first booster immunization and double booster immunization are shown in Supplementary Tables 1 and 2. Supplementary Table 1 also shows data for 8 November 2021, 31 December 2021, and 23 February 2022.

Vaccine effectiveness was examined for the predefined groups based on vaccination status versus the control, unvaccinated population during the Omicron and Delta waves. Additionally, registered rates of SARS-CoV-2 infection and Covid-19 related mortality were compared between the booster and double booster vaccinated populations.

Person-days for vaccination groups by wave were determined by summing the number of persons in a certain group for each day of the two periods from time zero (T0, 8 November 2021 and 1 January 2022, respectively). Average population sizes for all evaluated groups were calculated by dividing the total person-days of a given subgroup by the length of the study period (54 days). Data were stratified by age (16–24, 25–34, 35–44, 45–54, 55–64, 65–74, 75–84, 85≤ years).

Mortality rates were calculated by dividing the number of Covid-19 related deaths by the person-days of observation. Mortality rate ratios were estimated together with their exact 95% confidence intervals (95% CI) by age stratum. Mantel-Haenszel mortality rate ratios were estimated for the total populations to adjust for age.

The study was approved by the Central Ethical Committee of Hungary (OGYÉI/10296-1/2022 and IV/1722-1/2022/EKU) and followed the Strengthening the Reporting of Observational Studies in Epidemiology (STROBE) guidelines (17).

## 3 Results

Altogether 8,244,440 persons constituted the total study population during the Delta wave. 314,700 persons had SARS-CoV-2 infection (3.82%), of whom 6,571 persons died (0.080% of the investigated population) during the study period (Suppl Fig 1). The average size of the unvaccinated population (without prior SARS-CoV-2 infection) comprised 2,226,526 persons during the Delta wave (27.0% of total), of whom 3,834 (0.172%) died due to Covid-19 (Figure 2). In the total Delta population, the average size of the population with primary immunization was 3,476,436 (42.2%), of whom 2,071 (0.057%) died while having SARS-CoV-2 infection during this study period. 21.1% of this total population received booster immunization, of whom 0.022% (443 persons) died (Figure 2). Mortality rates increased by age, up to 30.38, 13.14, and 2.64 per 100.000 person-days in the oldest age cohort in the non-vaccinated, primary immunized, and booster vaccinated population, respectively.

**Figure 2.**
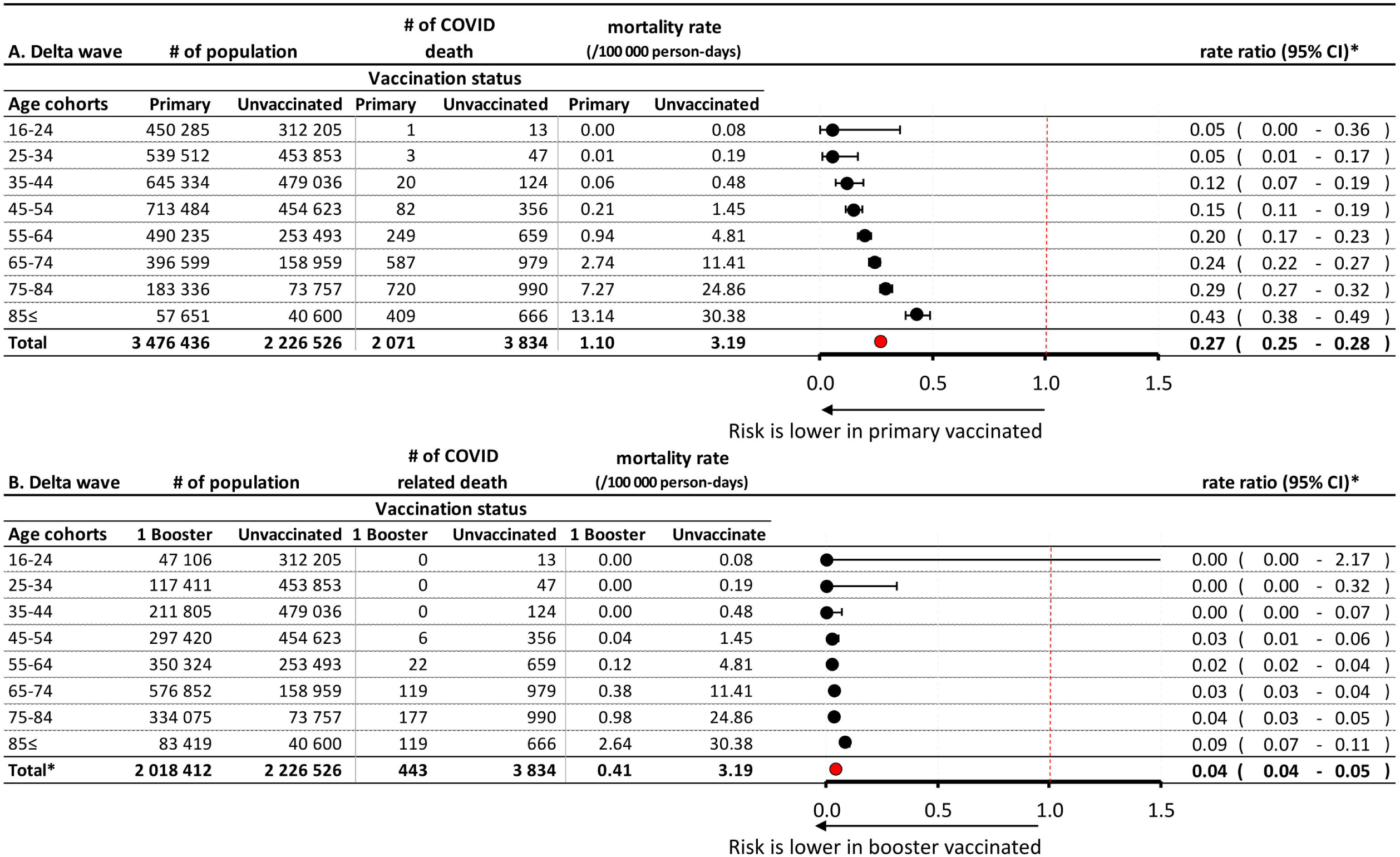
Risk of Covid-19 related mortality during the Delta wave in the primary immunized vs. unvaccinated control population (A), and in the booster vaccinated vs. unvaccinated control population (B), according to age. *Exact confidence intervals for age-specific mortality rate ratios and Mantel-Haenszel pooled mortality rate ratio for the total population adjusted for age.

The total study population for the period of the Omicron wave comprised 8,232,169 persons, of whom 2,792 (0.036%) died due to Covid-19. The average size of the primary immunized and booster vaccinated populations was 2,328,562 persons (28.3%) and 3,297,024 (40.05%), respectively (Suppl fig 1). Of the latter, 66.43% were aged 65 years or older. 31,527 persons (0.38%) had received double booster immunization (mostly a fourth dose, expect for the Janssen vaccine). In the booster vaccinated population, 845 persons (0.026%) died while having SARS-CoV-2 infection, while in the double booster immunized group, only 1 person died while having SARS-CoV2 infection during the Omicron wave (0.003%). Mortality rates were generally lower during the Omicron wave in most study groups compared to the Delta wave: 1.08 vs. 3.19 per 100.000 person-days in non-vaccinated cohorts, 0.48 vs. 1.10 per 100.000 person-days in the primary immunized populations, and 0.48 vs. 0.41 in the booster vaccinated populations, respectively. During the Omicron wave, the rate of Covid-19 related mortality was only 0.06 per 100.000 person-days in the double booster immunized group.

During the Delta wave, the relative rate (RR) of Covid-19 related mortality was 0.27 in the primary immunized population (95% CI: 0.25–0.28) compared to the unvaccinated control group. The rate ratio increased with age (0.05 in the 16–34 years, and 0.24–0.43 in the 65≤ years age groups). The risk of Covid-19 related mortality was 96% lower in the booster vaccinated population than in the unvaccinated control group (RR: 0.04; 95% CI: 0.04–0.05). The rate ratio was below 0.05% in most age groups, and there was a 100% risk reduction among people aged 0–45 years.

During the Omicron wave in 2022, people with primary immunization had a 40% lower risk of Covid-19 related death than those who received no immunization (RR: 0.60; 95% CI: 0.55–0.65) (Figure 3A). The difference decreased with age. The booster vaccinated population had an 82% lower risk of Covid-19 related mortality compared to unvaccinated controls (RR: 0.18; 95% CI: 0.16–0.20), with a slight decrease in effect with increasing age (Figure 3B). Double booster immunized people had an almost 100% Covid-19 related mortality reduction during the Omicron wave compared to the unvaccinated control population (RR: 0.01; 95% CI: 0.00–0.08) (Figure 3C).

**Figure 3.**
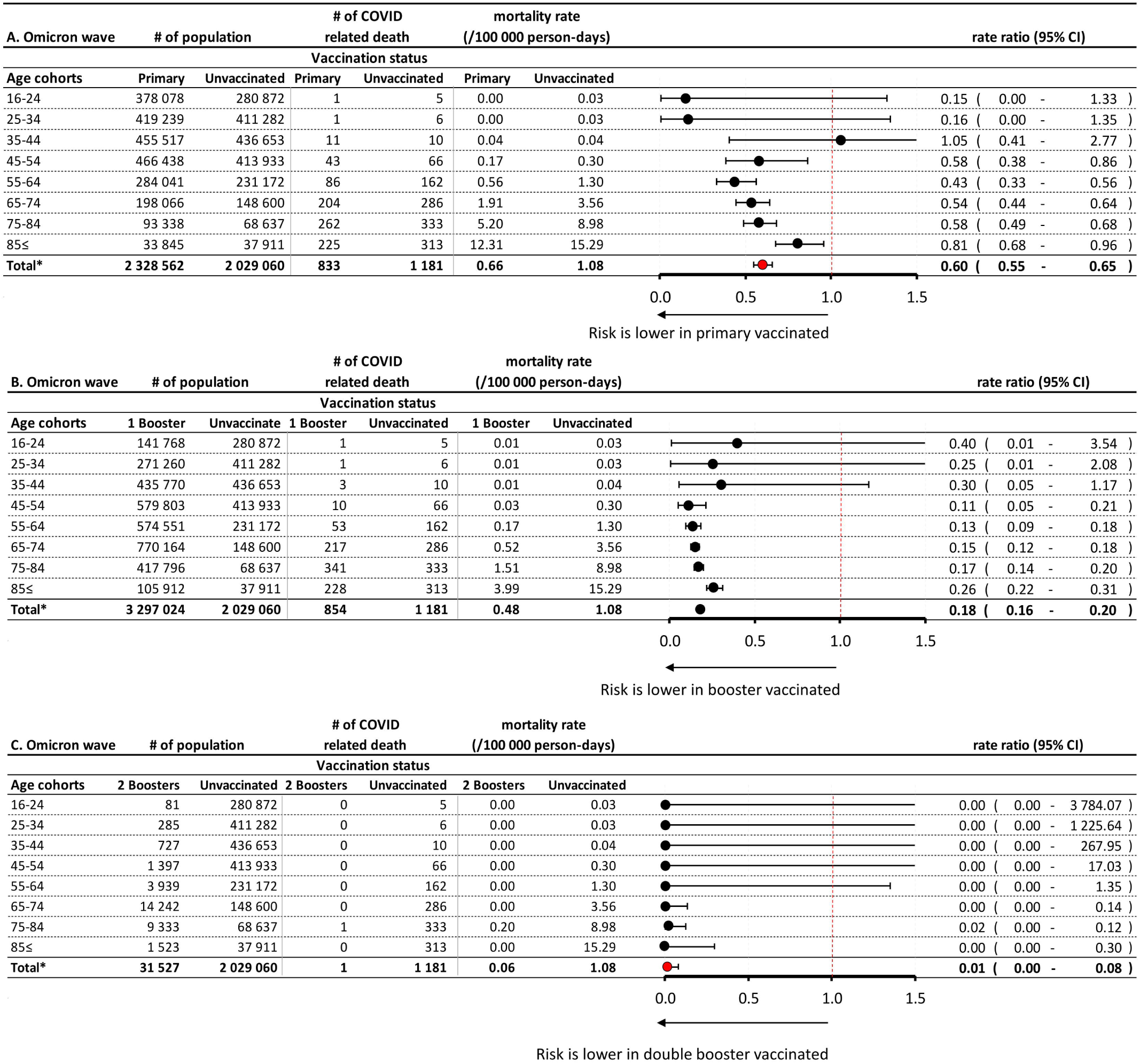
Risk of Covid-19 related mortality during the Omicron wave in the primary immunized vs. unvaccinated control population (A), in the booster vaccinated vs. unvaccinated control population (B), and in the double booster vaccinated vs. unvaccinated control population (C), according to age. *Exact confidence intervals for age-specific mortality rate ratios and Mantel-Haenszel pooled mortality rate ratio for the total population adjusted for age.

There was a 51% lower risk of registered SARS-CoV-2 infection and an almost 93% lower risk of Covid-19 related mortality among people with two booster doses compared to those with only one (RR: 0.49; 95% CI: 0.44–0.53 and RR: 0.07; 95% CI. 0.01–0.46, respectively) (Figure 4A–B). In the age group of 65≤ years, where the majority of booster doses were administered, the mortality reduction was statistically significant (i.e., the 95% CI of the mortality rate ratio does not include the value of 1 in the age groups 65–74 and 75–84 years).

**Figure 4.**
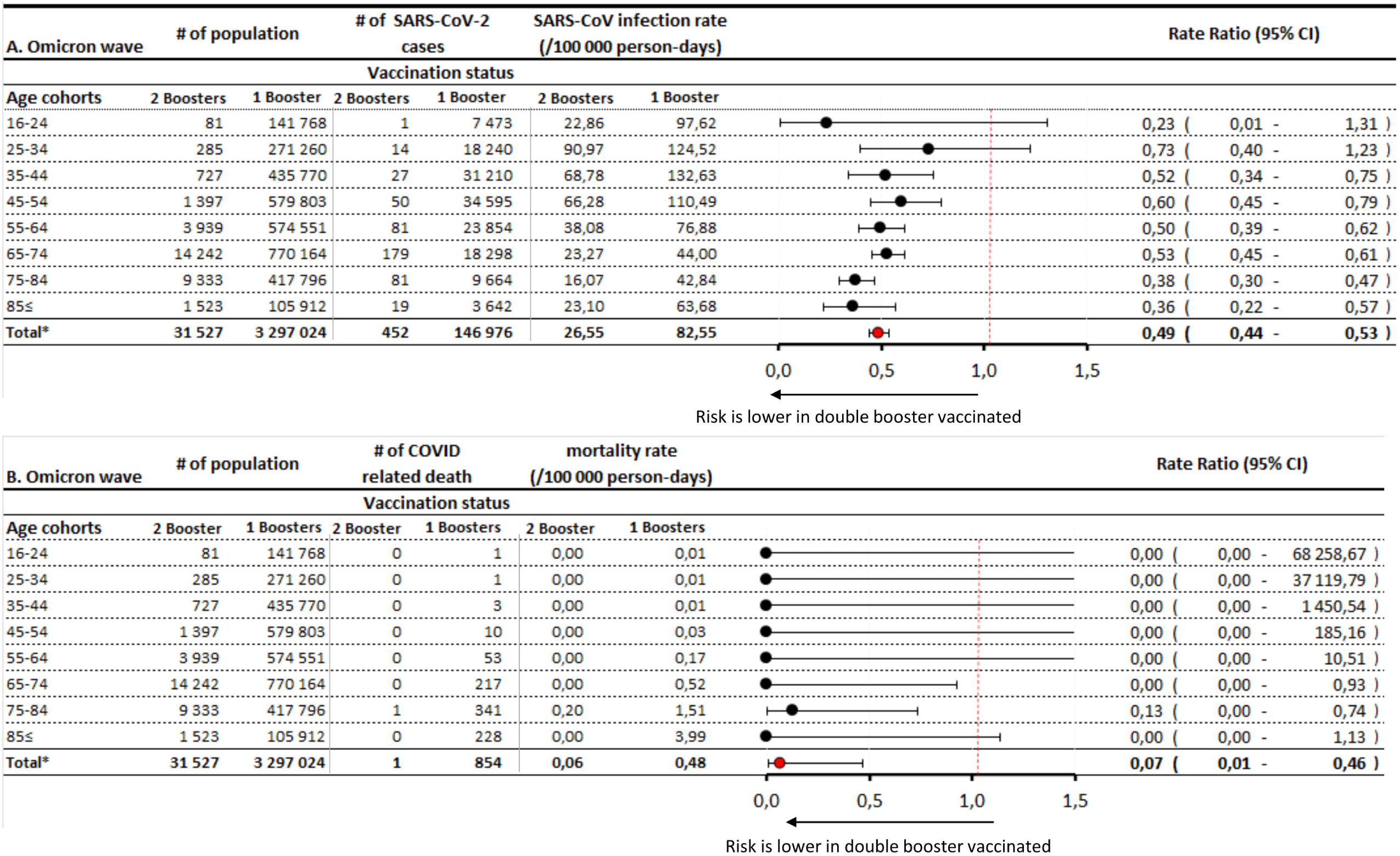
SARS-CoV-2 infection rates (A) and Covid-19 related mortality (B) during the Omicron wave in the double vs. single booster vaccinated population. *Exact confidence intervals for age-specific mortality rate ratios and Mantel-Haenszel pooled mortality rate ratio for the total population adjusted for age.

Overall, the risk of Covid-19 related mortality rate was 55% lower in the total Hungarian population during the Omicron vs. Delta wave (RR: 0.45 95%CI: 0.44-0.48). The difference decreased with age, with RRs of 0.15 (95% CI: 0.06–0.33) and 0.65 (95% CI: 0.59–0.71) in the age groups of 25–34 and 85≤ years, respectively (Supplementary Figure 1).

Booster vaccination provided an additional 85% reduction of mortality risk during the Delta wave (RR: 0.15; 95% CI: 0.13¬-0.17) and an additional 71% reduction during the second study period, dominated by the Omicron variant (RR: 0.29; 95% CI: 0.2–0.32) (Supplementary Figure 2A and 2B). This benefit had roughly the same size in almost all age cohorts.

## 4 Discussion

Our retrospective study aimed to estimate the effectiveness of primary and booster SARS-CoV-2 vaccination in the pandemic waves dominated by the Delta and Omicron variants in terms of Covid-19 related mortality and explore differences between the two waves in question.

Despite its higher transmissibility, the Omicron variant has been shown to be associated with a significantly lower risk of severe outcomes compared to the Delta variant (18,19). However, only few studies reported lower rates of the most severe outcome, Covid-19 related mortality. A recent study from South-Africa found a 73% lower risk of mortality with the Omicron vs. Delta variant (20), and the U.S. Centers for Disease Control and Prevention (CDC) reported mortality rates of 9/1,000 persons and 13/1,000 persons for the Omicron and Delta variants, respectively (21). In our study, the mortality rate of Covid-19 during the Omicron wave was 56% lower compared to the period dominated by the Delta wave in the total Hungarian population, with more pronounced differences in younger age groups. The lower risk of Covid-19 related mortality associated with the Omicron variant may be attributed to multiple factors including its potentially lower virulence (22,23), the increasing proportion of booster immunized people by the time of the Omicron wave, and the significant impact of detected and undetected previous Delta infections (24) which could have provided protection against severe outcomes during the Omicron wave (25). Our results demonstrate the lower risk of mortality associated with the Omicron variant and confirm previous observations from South Africa and the United States.

In our study, the risk of Covid-19 related mortality was 73% lower in people who had received primary immunization (approx. 3.5 million people) compared to the unvaccinated control group during the Delta wave. Booster vaccination, mainly with an mRNA vaccine, provided additional benefits with a 97% lower risk of Covid-19 related mortality compared to the unvaccinated population. The results show that although primary immunization (mainly administered in spring 2021) had already lost effectiveness against SARS-CoV-2 infection by the time of our study period, it still provided significant additional protection against Covid-19 related death (85% relative risk reduction) (26). Our observations are in line with a study conducted among 780,225 U.S. veterans which reported significantly declining vaccine effectiveness for all investigated vaccine types against symptomatic SARS-CoV-2 infection but sustained protection against Covid-19 related death among those infected during the Delta wave (52.2% for Janssen, 75.5% for Moderna, and 70.1% for Pfizer-BioNTech in the age group of ≥65 years) (5). Furthermore, several studies have demonstrated the additional benefits of booster immunization (9,10). In our study, this benefit was more apparent in older age groups. Compared to the unvaccinated control population, primary immunization and booster immunization resulted in a 76% and 97% lower risk of Covid-19 related mortality in the age group of 65–74 years, respectively.

The rationale for booster vaccines was questioned by several experts who argued that due to the high number of mutations affecting the spike protein of the Omicron variant (9), vaccine effectiveness against infection and severe outcomes may be compromised (21). In a study from the U.K., primary immunization with two doses of the Astra Zeneca or Pfizer-BioNTech vaccines provided limited or no protection against symptomatic disease caused by the Omicron variant, while an mRNA booster after the primary course of immunization significantly increased protection (67.2%–73.9%), although with waning effectiveness over time (60.9%–64.4%) (27).

In Hungary, the Omicron variant was first detected on 13 December 2021 (28) and became dominant by early January 2022 (29), reaching highest-ever daily infection rates on 31 January 2022. On 13 January 2022, the Hungarian government offered the option of a fourth dose for elderly people, healthcare workers and people with chronic diseases if at least 4 months had passed since their previous booster dose. By the end of our study period (23 February 2022), 189,894 people had received the second booster dose (although the average size of the population with at least 14 days since the second booster dose was only 31,527 during the study period), mostly mRNA vaccines (98.46%). There was a 40% lower risk of Covid-19 related mortality in the primary immunized population compared to the unvaccinated control group, despite higher infection rates in the former. In the booster immunized population, the risk of Covid-19 related death was 82% lower than in the unvaccinated group, and the few people who had received their second booster dose (n=31,527) had an almost 100% protection against mortality, with only one Covid-19 related death during the study period. The second booster dose further decreased the rate of SARS-CoV-2 infection by 51% compared to only one booster and provided an additional 93% mortality reduction. Although the short follow-up time of the double booster immunized group and the very low number of patients younger than 65 years in this group limit the interpretation of results related to the effectiveness of the second booster dose, these data confirm the additional short-term benefit of booster doses in the prevention of Covid-19 related mortality during the Omicron wave in the elderly.

The strengths of the HUN-VE-2 study include its nationwide nature, and the analysis of vaccine effectiveness during two consecutive waves of the coronavirus pandemic. Furthermore, the large study populations allowed for the evaluation of different levels of vaccination in different age groups. Nevertheless, our study has some limitations. The proportion of undetected cases might have been high and could have increased during the wave dominated by the Omicron variant. Therefore, we refrained from analyzing registered infections. We made an exemption when we estimated the impact of double booster and single booster vaccination, because we wanted to see how much the effect size is different in the case of SARS-CoV-2 infection and Covid-19 related mortality. Although the infection rates might be underestimated, we assume that the ascertainment bias was not likely to significantly affect the relative rates, as the proportion of unregistered cases among all cases was not likely to substantially differ in the two compared groups.

We studied the effect of vaccination by age, and adjusted for age when studying total populations, but an important limitation of our study is that we could not adjust for other potential confounders, most importantly for chronic diseases. As chronic diseases represent an indication for early vaccination as well as booster vaccination, and are closely related to mortality, this confounding may have influenced our results to a certain extent. However, as booster and second booster vaccinations were especially recommended for elderly people with chronic diseases, our estimates are conservative.

Our study is one of the very few demonstrating the benefit of second booster vaccination. Despite declining effectiveness against symptomatic disease caused by the Omicron variant, booster doses still provided protection against Covid-19 accompanying mortality. This suggests that although antibody responses tend to wane over time (27), cellular immune responses may also play a significant part in the protection against severe outcomes, and this protection may be significantly enhanced by a second booster dose. Our findings also raise the possibility that cellular immune responses may have an important role in the protection against upcoming new SARS-CoV-2 variants. A recent study from Israel demonstrated the rapid decline in the regained reduction of viral load achieved by the booster shot 3–4 months after its administration (30). These results strongly correlate with our findings showing more pronounced mortality risk reduction among people who received booster immunization during the Delta wave compared to the Omicron wave. Therefore, Hungarian guidelines recommend a fourth vaccine dose for the vulnerable population after 4 months of their first booster dose.

The nationwide HUN-VE 2 study provides an overview of the protection against Covid-19 related mortality with different levels of vaccination during the Delta and Omicron waves. The results show a significantly lower risk of Covid-19 related mortality during the Omicron wave in the whole study population and confirm the benefits of booster vaccination in the prevention of Covid-19 related death. Furthermore, our study is among the first to report the significant additional benefit of a second booster dose in terms of mortality reduction in older populations, beyond the improvement in protection against symptomatic SARS-CoV-2 infection.

## Data Availability

All data produced in the present study are available upon reasonable request to the authors

## 6 Conflict of Interest

*Zsófia Barcza of Syntesia Medical Communications Ltd. received payment for medical writing support from the National Public Health Center of Hungary. Zoltán Kiss is employed by MSD Pharma Hungary Ltd*., *too. However, this provides no relevant conflict of interest for the current research. At the time the study was performed, M*.*K served as the minister of human resources. The ministry includes the secretariat for health*.

## 7 Author Contributions

Z Vokó: Conceptualization, Methodology, Formal analysis, Validation and Writing – Review & Editing; Z Kiss and I Wittmann: Conceptualization, Methodology, Investigation, Visualization and Writing – Original Draft; L Polivka Conceptualization, Methodology, Formal analysis, Validation, Data curation; Gy Surján, P Nagy, I Kenessey, A Wéber: Conceptualization, Methodology, Validation, Review & Editing; O Surján, V Müller, Z Szekanecz, J Szlávik: Conceptualization, Review & Editing; M Kásler, C Müller, Zs Schaff, F Oberfrank, Gy Keserũ: Conceptualization, Methodology, Investigation and Supervision; Zs Barcza: Writing – Original Draft; GA Molnár: Validation, Project Administration

## 8 Funding

The Center for Health Technology Assessment of the Semmelweis University participated in the project on a contractual basis made with the Ministry of Human Resources of Hungary and received funding. Zsófia Barcza of Syntesia Medical Communications Ltd. received payment for medical writing support from the National Public Health Center of Hungary.

## 9 Acknowledgments

The authors would like to thank Richárd Kiss for the initial data curation and dataset validation.

## 11 Data Availability Statement

The datasets generated for this study can be found in the MedRxiv repository. Further inquiries can be directed to the corresponding author.

## 12 Contribution to the field statement

The Covid-19 pandemics led to an immense worldwide burden on healthcare systems, therefore development of vaccines was of high importance. The vaccines were tested in clinical trials against the original strain. However, with time, novel variants such as the alpha, delta and omicron have been arising. The efficacy of Covid-19 vaccines seems to wean over time; therefore, booster vaccines are frequently applied. We report on nationwide efficacy of vaccines in the Hungarian population. We compared the efficacy of first boostered vs. primary vaccinated vs. unvaccinated populations during both Delta and Omicron waves. Moreover, at the time of the Omicron wave, a second booster shot has been made available by the Hungarian government, thus we could also test double booster vaccinated vs. single booster vaccinated populations. Mortality due to Covid-19 was tested, and we found that primary vaccinated persons were more protected than unvaccinated ones during both waves. The first booster led to an increased efficacy. During the Omicron wave, the second booster provided and additional protection even when compared to the first booster-vaccinated population. Our study provides nationwide data on vaccine efficacy against two variants. As second booster was made available early, data are robust due to a large population.

## Notes

### Competing Interest Statement

Zsofia Barcza of Syntesia Medical Communications Ltd. received payment for medical writing support from the National Public Health Center of Hungary. Zoltan Kiss is employed by MSD Pharma Hungary Ltd., too. However, this provides no relevant conflict of interest for the current research. At the time the study was performed, M.K served as the minister of human resources. The ministry includes the secretariat for health.

### Author Declarations

The study was approved by the Central Ethical Committee of Hungary (OGYEI/10296-1/2022 and IV/1722- 1/2022/EKU) and followed the Strengthening the Reporting of Observational Studies in Epidemiology (STROBE) guidelines.

