## Supplementary Tables and Figures for "Nationwide Effectiveness of First and Second SARS-CoV2 Booster Vaccines during the Delta and Omicron Pandemic Waves in Hungary (HUN-VE 2 Study)"

Supplementary Material

### 1 Supplementary Figures

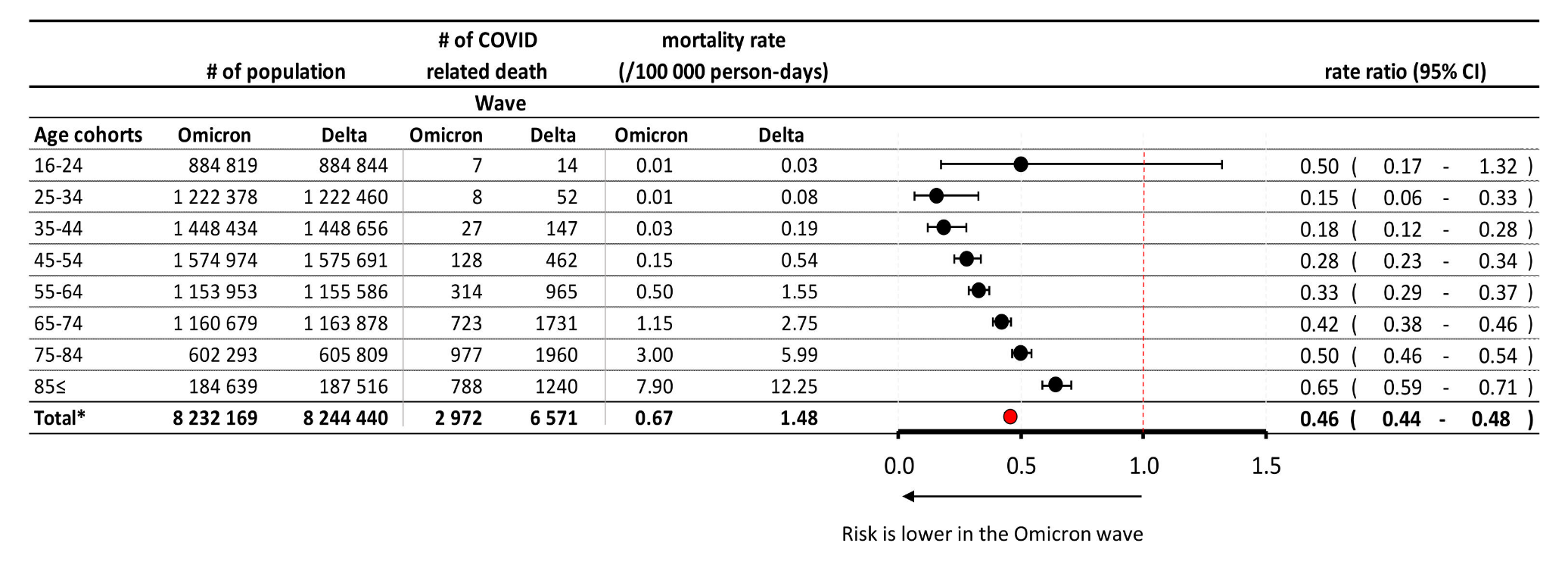

**Supplementary Figure 1.** Risk of Covid-19 related mortality in the whole study population during the Omicron vs. Delta wave, according to age. *Exact confidence intervals for age-specific mortality rate ratios and Mantel-Haenszel pooled mortality rate ratio for the total population adjusted for age.

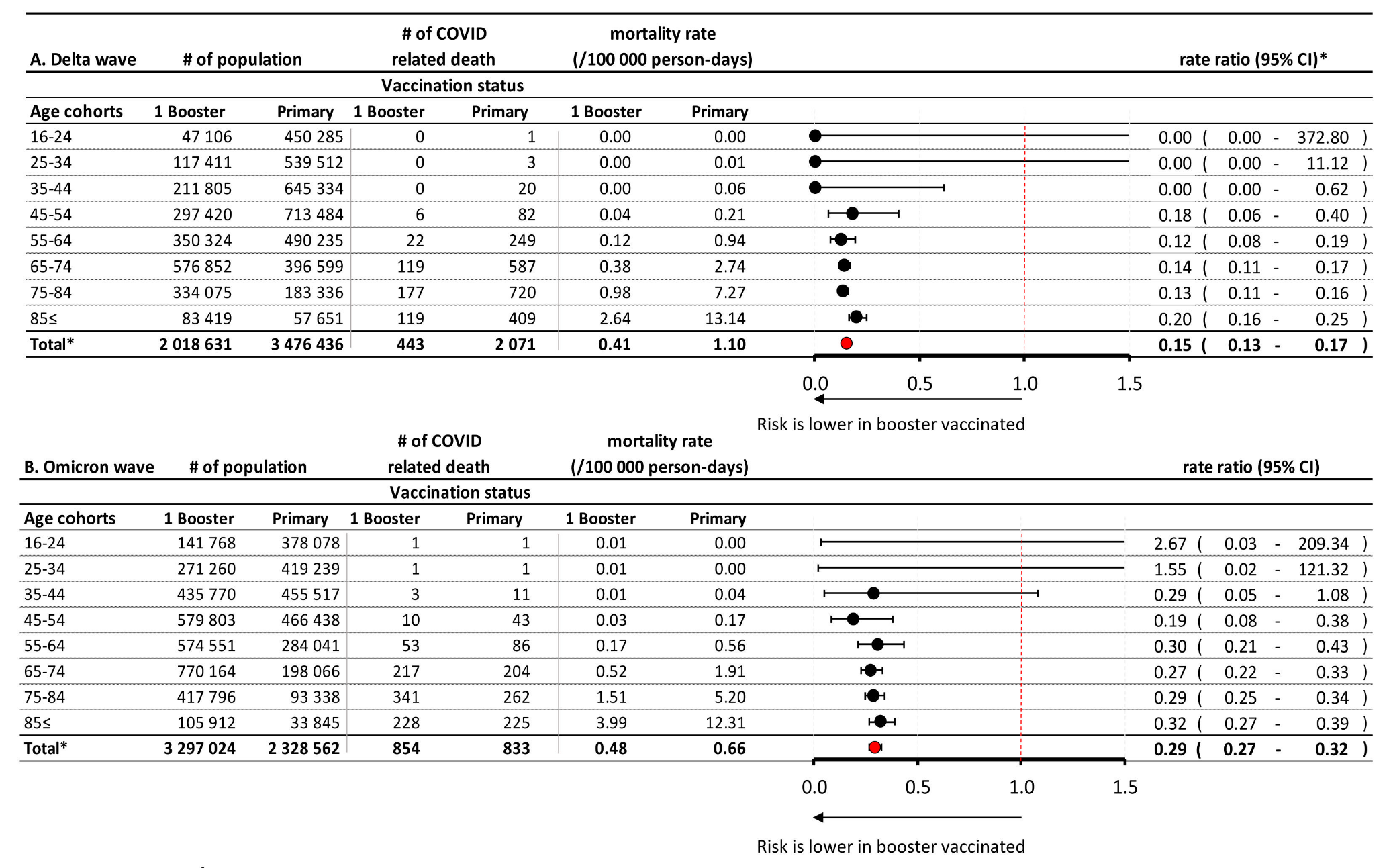
**Supplementary Figure 2.** Age-dependent risk of Covid-19 related mortality during the Delta wave (A) and during the Omicron wave (B) in the booster vaccinated vs. primary immunized population. *Exact confidence intervals for age-specific mortality rate ratios and Mantel-Haenszel pooled mortality rate ratio for the total population adjusted for age.

### 2 Supplementary Tables

**Supplementary Table 1.** Vaccination status at the beginning and end of the two study periods

| **Vaccination status  on 8 November 2021** | | **Only  1 vaccine administered** | **Complete primary vaccination** | **1st booster vaccination** | | | | | | | **Total vaccinated** |
| --- | --- | --- | --- | --- | --- | --- | --- | --- | --- | --- | --- |
|  |  |  |  | **Pfizer-BioNTech** | **Moderna** | **Sputnik-V** | **Astra Zeneca** | **Sinopharm** | **Jansen** | **Total boostered** |  |
| **Primary vaccination** | **Pfizer-BioNTech** | 125 123 | 2 149 762 | 367 855 | 10 454 | 340 | 1 474 | 11 719 | 15 188 | 407 030 | 2 681 915 |
|  | **Moderna** | 11 322 | 285 189 | 19 637 | 19 591 | 30 | 218 | 2 240 | 3 353 | 45 069 | 341 580 |
|  | **Sputnik-V** | 13 119 | 729 211 | 139 335 | 12 128 | 141 | 28 | 1 186 | 5 947 | 158 765 | 901 095 |
|  | **Astra Zeneca** | 14 735 | 509 165 | 81 041 | 7 512 | 10 | 185 | 712 | 349 | 89 809 | 613 709 |
|  | **Sinopharm** | 31 013 | 606 909 | 367 041 | 18 439 | 162 | 381 | 5 363 | 16 461 | 407 847 | 1 045 769 |
|  | **Janssen** | NA | 131 276 | 1 415 | 171 | - | 1 | 28 | 130 | 1 745 | 133 021 |
|  | **Heterologous primary immunization** | NA | 26 291 | 1 911 | 20 | 2 | 6 | 42 | 87 | 2 248 | 28 539 |
| **Total** | | 195 312 | 4 437 803 | 978 235 | 68 495 | 685 | 2 293 | 21 290 | 41 515 | 1 112 513 | 5 717 089 |
| **Vaccination status  on 31 December 2021** | |  |  |  |  |  |  |  |  |  |  |
| **Primary vaccination** | **Pfizer-BioNTech** | 194 789 | 1 413 808 | 1 156 690 | 37 252 | 379 | 1 969 | 24 185 | 30 218 | 1 250 693 | 2 859 290 |
|  | **Moderna** | 14 154 | 167 278 | 57 861 | 96 014 | 35 | 363 | 4 811 | 7 372 | 166 456 | 347 888 |
|  | **Sputnik-V** | 9 494 | 347 060 | 465 920 | 47 833 | 163 | 49 | 6 733 | 19 583 | 540 281 | 896 835 |
|  | **Astra Zeneca** | 9 482 | 203 766 | 348 847 | 39 793 | 19 | 1 853 | 3 350 | 1 282 | 395 144 | 608 392 |
|  | **Sinopharm** | 43 431 | 365 270 | 567 611 | 36 093 | 178 | 488 | 23 074 | 31 162 | 658 606 | 1 067 307 |
|  | **Janssen** | NA | 157 535 | 14 542 | 1 977 | 1 | 3 | 367 | 3 260 | 20 150 | 177 685 |
|  | **Heterologous primary immunization** | NA | 32 921 | 7 966 | 923 | 2 | 21 | 212 | 213 | 9 337 | 42 258 |
| **Total** | | 271 350 | 2 687 638 | 2 619 437 | 259 885 | 777 | 4 746 | 62 732 | 93 090 | 3 040 667 | 5 999 655 |
| **Vaccination status on 23 February 2022** | |  |  |  |  |  |  |  |  |  |  |
| **Primary vaccination** | **Pfizer-BioNTech** | 139 265 | 1 234 546 | 1 410 544 | 45 741 | 309 | 1 699 | 26 824 | 30 587 | 1 515 704 | 3 064 056 |
|  | **Moderna** | 10 466 | 128 629 | 72 118 | 119 068 | 29 | 338 | 5 670 | 7 680 | 204 903 | 356 559 |
|  | **Sputnik-V** | 8 306 | 257 880 | 524 676 | 55 309 | 159 | 49 | 9 583 | 21 524 | 611 300 | 877 496 |
|  | **Astra Zeneca** | 8 113 | 125 357 | 409 102 | 47 682 | 23 | 2 203 | 4 693 | 1 625 | 465 328 | 598 847 |
|  | **Sinopharm** | 31 470 | 303 606 | 533 519 | 37 588 | 136 | 390 | 36 145 | 31 737 | 639 515 | 976 196 |
|  | **Janssen** | NA | 148 405 | 28 004 | 3 604 | 5 | 5 | 838 | 8 163 | 40 619 | 190 297 |
|  | **Heterologous primary immunization** | NA | 34 084 | 11 130 | 1 219 | 2 | 26 | 386 | 256 | 13 019 | 47 103 |
| **Total** | | 197 620 | 2 232 507 | 2 989 093 | 310 211 | 663 | 4 710 | 84 139 | 101 572 | 3 490 388 | 6 110 554 |

**Supplementary Table 2.** 2^nd^ booster vaccination status on 23 February 2022.

| **Vaccination stats  on 23 February 2022** | **2nd booster (regardless of 1st-3rd)** |
| --- | --- |
| Pfizer-BioNTech | 174 541 |
| Moderna | 12 561 |
| Sputnik-V | 10 |
| Astra Zeneca | 49 |
| Sinopharm | 1 605 |
| Jansen | 1 273 |
| **Total** | **190 039** |
